# Neighborhood Disadvantage and Active, Disabled and Total Life Expectancy Among Community-living Older Persons

**DOI:** 10.1101/2021.02.05.21251217

**Authors:** Thomas M. Gill, Emma X. Zang, Terrence E. Murphy, Linda Leo-Summers, Evelyne A. Gahbauer, Natalia Festa, Jason R. Falvey, Ling Han

## Abstract

**Background:** Neighborhood disadvantage is a novel social determinant of health that could adversely affect the functional well-being and longevity of older persons. We evaluated whether estimates of active, disabled and total life expectancy differ on the basis of neighborhood disadvantage after accounting for individual-level socioeconomic characteristics and other prognostic factors.

**Methods:** We used data on 754 community-living older persons from South Central Connecticut, who completed monthly assessments of disability from 1998 to 2020. Scores on the area deprivation index were dichotomized at the 80^th^ state percentile to distinguish neighborhoods that were disadvantaged (81-100) from those that were not (1-80).

**Results:** Within 5-year age increments from 70 to 90, active and total life expectancy were consistently lower in participants from neighborhoods that were disadvantaged versus not disadvantaged, and these differences persisted and remained statistically significant after adjustment for individual-level race/ethnicity, education, income, and other prognostic factors. At age 70, adjusted estimates (95% CI) for active and total life expectancy (in years) were 12.3 (11.5-13.1) and 15.0 (13.8-16.1) in the disadvantaged group and 14.2 (13.5-14.7) and 16.7 (15.9-17.5) in the non-disadvantaged group. At each age, participants from disadvantaged neighborhoods spent a greater percentage of their projected remaining life disabled, relative to those from non-disadvantaged neighborhoods, with adjusted values (SE) ranging from 17.7 (0.8) vs. 15.3 (0.5) at age 70 to 55.0 (1.7) vs. 48.1 (1.3) at age 90.

**Conclusions:** Living in a disadvantaged neighborhood is associated with lower active and total life expectancy and a greater percentage of projected remaining life disabled.

---

Neighborhood disadvantage is a novel social determinant of health that could adversely affect the functional well-being and longevity of older persons. A neighborhood may be disadvantaged based on indicators of education, employment, housing-quality and poverty. Prior research has shown that living in a socioeconomically disadvantaged neighborhood confers increased risk for several adverse outcomes including all-cause mortality,^1^ hospital readmission,^2^ the incidence and severity of delirium after major surgery,^3^ and Alzheimer disease neuropathology.^4^

Active and disabled life expectancy, defined as the projected number of remaining years without and with disability in activities of daily living, are often used by policy-makers to forecast the functional well-being of older persons.^5,6^ Whether these longitudinal metrics of functional well-being are influenced by neighborhood disadvantage is unknown. In a prior study of Medicare Advantage recipients, neighborhood disadvantage was associated cross-sectionally with difficulty in activities of daily living,^7^ but longitudinal data are lacking.

In the current study, we evaluated whether estimates of active, disabled and total life expectancy differ on the basis of neighborhood disadvantage. Because disadvantaged neighborhoods disproportionately include racial and ethnic minorities and persons of low socioeconomic status, we also evaluated whether these differences persist after accounting for individual-level socioeconomic characteristics and other prognostic factors. To accomplish our objectives, we used data from a longitudinal study that includes monthly assessments of disability in activities of daily living for more than 20 years in a cohort of community-living older persons, with little missing data and few losses to follow-up. The results from this study could inform policies to improve the functional well-being and longevity of older persons.

## METHODS

### Study Population

Participants were members of the Precipitating Events Project, a longitudinal study of 754 community-living persons, aged 70 years or older, who were nondisabled (i.e. required no personal assistance) in four essential activities of daily living—bathing, dressing, walking inside the house, and transferring from a chair. The assembly of the cohort, which took place between March 1998 and October 1999, has been described in detail elsewhere.^8,9^ In brief, potential participants were identified from a computerized list of 3,157 age-eligible members of a large health plan in South Central Connecticut. Eligibility was determined during a screening telephone interview and was confirmed during an in-home assessment. Persons were excluded based on the following criteria: significant cognitive impairment with no available proxy,^10^ inability to speak English, diagnosis of a terminal illness with a life expectancy less than 12 months, and plan to move out of the area during the next 12 months. Only 4.6% of the 2,753 health plan members who were alive and could be contacted refused to complete the screening telephone interview, and 75.2% of the eligible members agreed to participate in the project, which was approved by the Human Investigation Committee at Yale University. Persons who declined to participate did not differ from those who were enrolled in terms of age or sex.

### Data Collection

Comprehensive assessments were completed at baseline by trained nurse researchers, while telephone interviews were completed each month by a separate team of researchers through June 2020. For participants who had significant cognitive impairment or were otherwise unavailable, a proxy informant was interviewed using a rigorous protocol.^10^ Deaths were ascertained by review of local obituaries and/or from an informant during a subsequent telephone interview. Six hundred seventy-one (89.0%) participants died after a median of 110 months, while 47 (6.2%) withdrew from the study after a median of 26 months. Data were otherwise available for 99.2% of 86,689 monthly interviews. The cohort has been linked to Medicare data.^11^

#### Descriptive Characteristics and Covariates

During the baseline assessment, data were collected on demographic characteristics, years of education, income, chronic conditions, and cognition. Based on a card with 13 income categories, participants were asked, “Which of these groups represents your (and your spouse’s) income for the past year? Include income from all sources such as wages, salaries, social security or retirement benefits, help from relatives, rent from property and so forth.” Responses were subsequently classified into three income groups, with the lowest category (<$10,000) corresponding to the 1998 federal poverty threshold from the Census Bureau.^12^ The 9 self-reported, physician-diagnosed chronic conditions included hypertension, myocardial infarction, congestive heart failure, stroke, diabetes mellitus, arthritis, hip fracture, chronic lung disease, and cancer (other than minor skin cancers). Cognition was assessed with the Mini-Mental State Examination, with a score less than 24 denoting cognitive impairment.^13^

Collection of these baseline data was 100% complete with the exception of income, which was missing (refused or don’t know) for 51 (6.8%) participants. These missing values were imputed using data from 25 baseline covariates.^14,15^

Medicaid eligibility was ascertained through linkages with Medicare data.^11^

#### Neighborhood Disadvantage

Neighborhood disadvantage was assessed with the area deprivation index (ADI), a census-based socioeconomic index.^2,16^ The ADI includes 17 education, employment, housing quality, and poverty indicators obtained from the American Community Survey (**Table S1**). These indicators are weighted and summed to yield a score for each neighborhood at the census-block group level.^1^ For the current study, 9-digit zip codes from the Medicare Master Beneficiary Summary File (MBSF) were linked to the 2000 ADI scores and national percentiles using the Neighborhood Atlas.^2^ When not available from the MBSF, zip codes were determined from home addresses at baseline. The ADI scores were ranked to determine the state percentile for each participant. Higher ADI percentile scores indicate greater socioeconomic disadvantage. Based on its established threshold effects,^2^ the ADI scores were dichotomized at the 80^th^ state percentile to distinguish the disadvantaged (81-100) from the non-disadvantaged groups (1-80). State percentiles were used because policies most relevant to life expectancy are established at the state rather than national level.^17^ Because Connecticut is more affluent than the US as a whole, the corresponding national percentiles for the disadvantaged group were 46 or higher.

#### Monthly telephone interviews

Complete details regarding the assessment of disability, including reliability and accuracy, are provided elsewhere.^10,18^ Each month, participants were asked, “At the present time, do you need help from another person to [complete the task]?” for each of the four essential activities of daily living that were assessed during the screening telephone interview.^10^ Participants who needed help with any of the tasks were considered to be disabled. Conversely, those who did not need help were considered to be nondisabled (or independent). Participants were not asked about eating, toileting, or grooming. The incidence of disability in these three activities of daily living is low among nondisabled, community-living older persons.^19,20^ Furthermore, it is highly uncommon for disability to develop in these activities of daily living without concurrent disability in bathing, dressing, walking, or transferring.^19-21^ To address the small amount of missing data on disability, multiple imputation was used with 100 random draws per missing observation.^22^

### Statistical Analysis

Baseline characteristics were compared between the disadvantaged and non-disadvantaged groups. Active and disabled life expectancy were estimated using multi-state life tables under the assumption of a discrete-time Markov process.^23^ Each participant’s monthly probabilities of transitioning from their current state (active or disabled) to one of three states (active, disabled or death) were determined using a multinomial logistic model, with death as an absorbing state and age in months as an explanatory variable. Values for active, disabled, and total life expectancy were estimated separately for the disadvantaged and non-disadvantaged groups and are reported by age in 5-year increments from 70 to 90. For each age-specific set of values, 95% confidence intervals were calculated using 1000 bootstrap samples.^23,24^ The adjusted analyses included sex, race/ethnicity, years of education, and baseline values for living alone, income, Medicaid eligibility, number of chronic conditions, and cognitive impairment. For comparisons between the two groups, differences were considered to be statistically significant if the corresponding confidence intervals did not overlap.

For both the unadjusted and adjusted results, we calculated absolute and relative differences in active life expectancy between the disadvantaged and non-disadvantaged groups according to age and the percentage of projected remaining life with disability according to age and neighborhood disadvantage. For each set of values, standard errors were calculated using 1000 bootstrap samples.^23,24^

SAS version 9.2 (SAS Institute, Cary, NC) was used for the descriptive analyses, Stata Release 16 (StataCorp. 2019. College Station, TX: StataCorp LLC) was used for the multinomial logistic regression analyses, and R code in the RStudio environment (RStudio Version 1.2.1335 © 2009-2019 RStudio, Inc.) was used for the bootstrapped estimates.

## RESULTS

The baseline characteristics of the study population are provided in **Table 1**. Compared with participants who were living in neighborhoods that were not disadvantaged, participants in the disadvantaged group were more likely to be female, Medicaid eligible, and cognitively impaired, but were less likely to be Non-Hispanic white and to live alone. They also had fewer years of education and lower income. The small differences in age and number of chronic conditions between the two groups were not statistically significant.

**Table 1.**
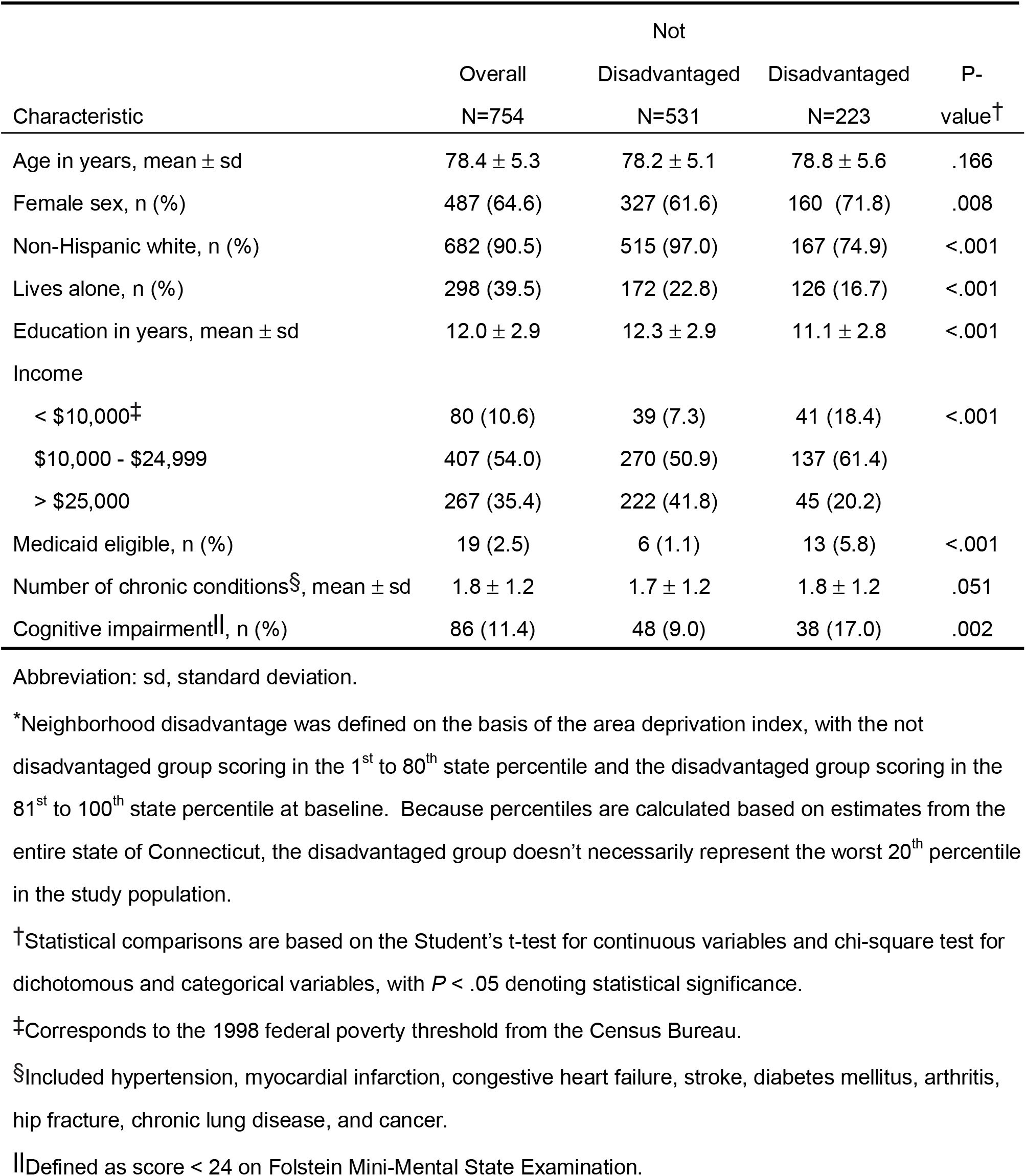
Baseline Characteristics According to Neighborhood Disadvantage*

Among all participants, mean estimates (95% confidence interval) for active, disabled, and total life expectancy, anchored at age 70, were 13.2 (12.6, 13.7), 2.7 (2.5, 2.9), and 15.8 (15.1, 16.5) years, respectively. **Table 2** provides these values according to age and neighborhood disadvantage. In both the unadjusted and adjusted analyses, active life expectancy was significantly lower in participants from disadvantaged neighborhoods than those from non-disadvantaged neighborhoods at each age. As shown in **Figure 1**, absolute differences were larger at the younger ages, while relative differences were larger at the older ages. These differences were only slightly attenuated in the adjusted analyses. In contrast, while disabled life expectancy was consistently higher in participants from disadvantaged neighborhoods in the unadjusted analyses, these non-significant differences were diminished in the adjusted analyses, largely due to reductions in disabled life expectancy among participants in the disadvantaged group. For participants with and without neighborhood disadvantage, the adjusted values for disabled life expectancy were comparable across the different ages, ranging from 2.4 to 2.7 years for participants from disadvantaged neighborhoods and 2.3 to 2.6 years for participants from non-disadvantaged neighborhoods. Differences in total life expectancy tended to track most closely to those in active life expectancy, with values being consistently lower among participants from disadvantaged neighborhoods than those from non-disadvantaged neighborhoods in both the unadjusted and adjusted analyses.

**Table 2.** Active, Disabled and Total Life Expectancy According to Age and Neighborhood Disadvantage

|  |  | Unadjusted Model |  |  |  |  |  | Adjusted Model<sup>†</sup> |  |  |  |  |  |
| --- | --- | --- | --- | --- | --- | --- | --- | --- | --- | --- | --- | --- | --- |
|  |  | Active Life |  | Disabled Life |  | Total Life |  | Active Life |  | Disabled Life |  | Total Life |  |
|  |  | Expectancy, y |  | Expectancy, y |  | Expectancy, y |  | Expectancy, y |  | Expectancy, y |  | Expectancy, y |  |
| Age, y | Disad-<br>vantaged* | Mean | 95% CI | Mean | 95% CI | Mean | 95% CI | Mean | 95% CI | Mean | 95% CI | Mean | 95% CI |
| 70 | No | 13.8 | 13.2, 14.4 | 2.5 | 2.3, 2.7 | 16.3 | 15.5, 17.1 | 14.2 | 13.5, 14.7 | 2.6 | 2.3, 2.8 | 16.7 | 15.9, 17.5 |
|  | Yes | 11.7 | 10.9, 12.5 | 3.0 | 2.6, 3.5 | 14.7 | 13.6, 15.9 | 12.3 | 11.5, 13.1 | 2.7 | 2.3, 3.1 | 15.0 | 13.8, 16.1 |
| 75 | No | 10.0 | 9.6, 10.5 | 2.5 | 2.3, 2.7 | 12.5 | 11.9, 13.1 | 10.2 | 9.8, 10.7 | 2.5 | 2.3, 2.8 | 12.7 | 12.1, 13.4 |
|  | Yes | 8.2 | 7.7, 8.8 | 3.0 | 2.6, 3.4 | 11.2 | 10.3, 12.1 | 8.6 | 8.0, 9.2 | 2.6 | 2.2, 3.0 | 11.2 | 10.3, 12.2 |
| 80 | No | 6.8 | 6.5, 7.1 | 2.5 | 2.3, 2.7 | 9.3 | 8.8, 9.8 | 6.9 | 6.6, 7.2 | 2.5 | 2.3, 2.7 | 9.3 | 8.9, 9.9 |
|  | Yes | 5.4 | 5.0, 5.8 | 2.9 | 2.6, 3.4 | 8.3 | 7.6, 9.1 | 5.6 | 5.2, 6.0 | 2.6 | 2.2, 2.9 | 8.1 | 7.4, 8.9 |
| 85 | No | 4.3 | 4.1, 4.6 | 2.4 | 2.2, 2.7 | 6.7 | 6.3, 7.2 | 4.3 | 4.0, 4.5 | 2.4 | 2.2, 2.7 | 6.7 | 6.2, 7.1 |
|  | Yes | 3.3 | 3.0, 3.6 | 2.9 | 2.5, 3.3 | 6.2 | 5.6, 6.8 | 3.4 | 3.1, 3.7 | 2.5 | 2.1, 2.9 | 5.8 | 5.3, 6.5 |
| 90 | No | 2.6 | 2.4, 2.8 | 2.4 | 2.1, 2.7 | 4.9 | 4.5, 5.4 | 2.5 | 2.3, 2.7 | 2.3 | 2.0, 2.6 | 4.8 | 4.4, 5.2 |
|  | Yes | 1.9 | 1.7, 2.1 | 2.8 | 2.4, 3.3 | 4.8 | 4.2, 5.4 | 1.9 | 1.7, 2.1 | 2.4 | 2.0, 2.8 | 4.3 | 3.8, 4.9 |

Abbreviation: CI, confidence interval.
\*Denotes neighborhoods in the 81st to 100th state percentiles of the area deprivation index at baseline.
<sup>†</sup>Includes sex, race/ethnicity, years of education, and baseline values for living alone, income, Medicaid eligibility, number of chronic conditions, and cognitive impairment.

**Figure 1.**
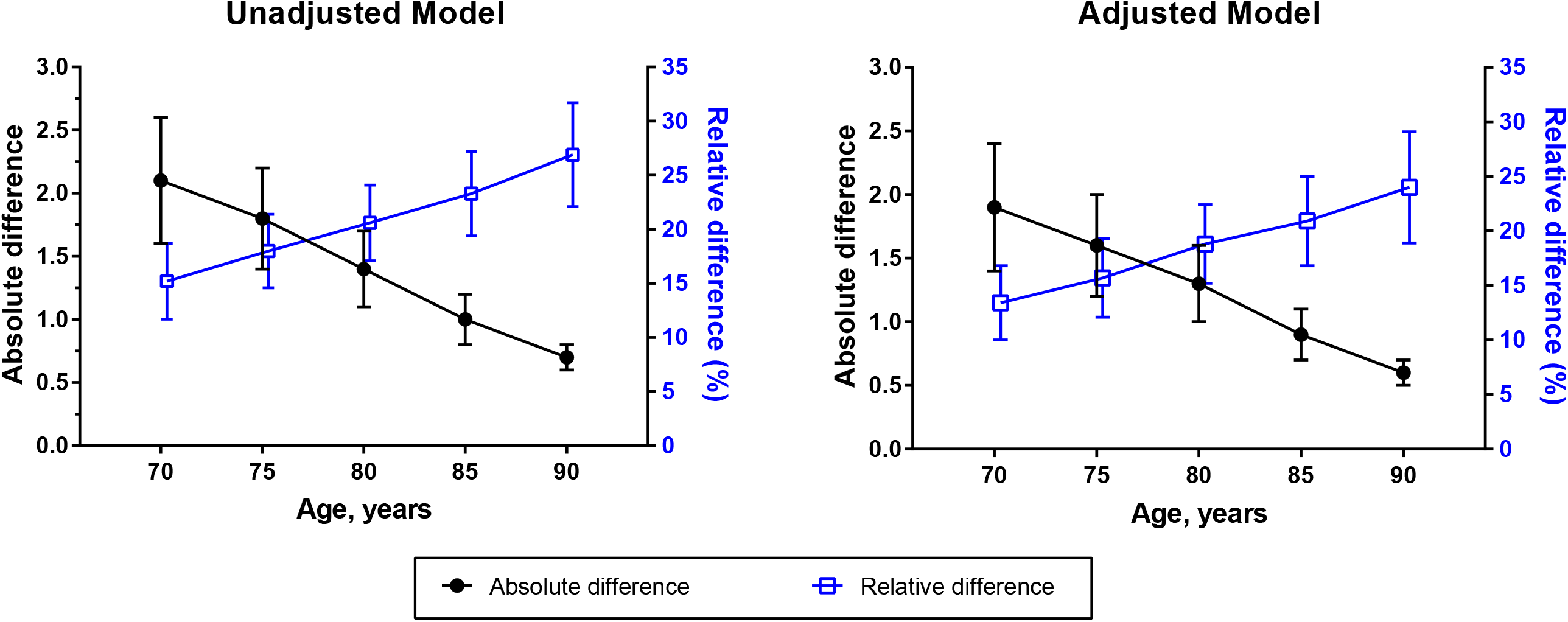
Absolute and Relative Differences in Active Life Expectancy Between Participants Living in Neighborhoods that were Not Disadvantaged Versus Disadvantaged According to Age. Values are accompanied by standard error bars. Neighborhoods in the 81st to 100th state percentiles of the area deprivation index were classified as disadvantaged. The denominators for the relative differences are based on the active life expectancies of participants who were not disadvantaged. The adjusted models included sex, race/ethnicity, years of education, and baseline values for living alone, income, Medicaid eligibility, number of chronic conditions, and cognitive impairment.

Values for the percentage of projected remaining life with disability are provided in **Figure 2**. In both the unadjusted and adjusted analyses, values increased progressively with advancing age for participants with and without neighborhood disadvantage. At each age, however, participants from disadvantaged neighborhoods spent a greater percentage of their projected remaining life with disability, relative to those from non-disadvantaged neighborhoods, and these differences were only modestly diminished in the adjusted analyses.

**Figure 2.**
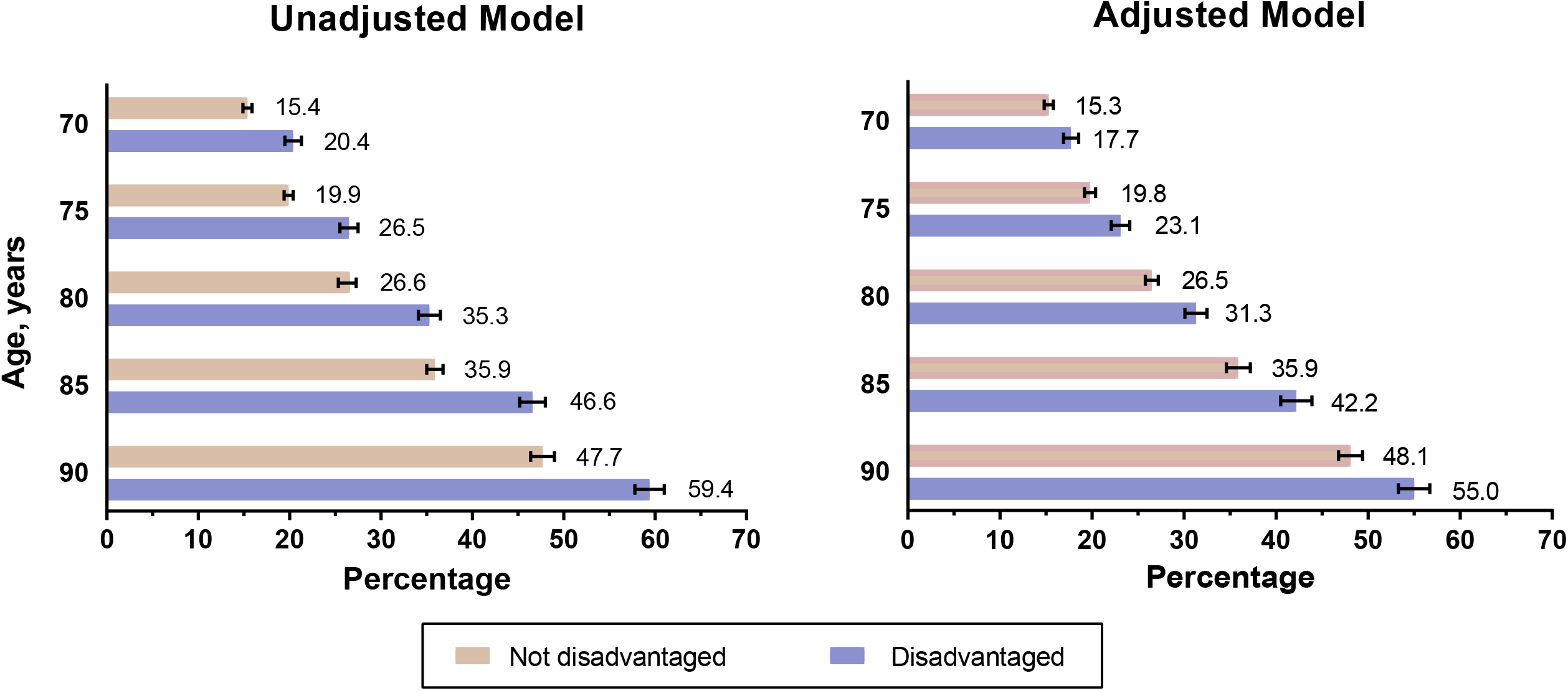
The Percentage of Projected Remaining Life with Disability According to Age and Neighborhood Disadvantage. Values are accompanied by standard error bars. Neighborhoods in the 81st to 100th state percentiles of the area deprivation index were classified as disadvantaged. The adjusted models included sex, race/ethnicity, years of education, and baseline values for living alone, income, Medicaid eligibility, number of chronic conditions, and cognitive impairment.

## DISCUSSION

In this prospective longitudinal study of nondisabled community-living older persons, we evaluated whether estimates of active, disabled and total life expectancy differ on the basis of neighborhood disadvantage within 5-year age increments from 70 to 90. Three major findings warrant comment. First, active life expectancy and total life expectancy were consistently lower in participants who were living in neighborhoods that were disadvantaged relative to those who were living in neighborhoods that were not disadvantaged, and these differences persisted and remained statistically significant after adjustment for individual-level socioeconomic characteristics and other prognostic factors. Second, disabled life expectancy did not differ significantly between participants with and without neighborhood disadvantage. Third, participants from disadvantaged neighborhoods spent a greater percentage of their projected remaining life with disability, relative to those from neighborhoods that were not disadvantaged. These findings suggest that living in a socioeconomically disadvantaged neighborhood may adversely affect the functional well-being and longevity of older persons.

Several potential mechanisms may underlie our findings. First, participants living in resource-poor environments may have less ready access to high-quality medical care,^25^ leading to less aggressive efforts to prevent, manage, and remediate episodes of functional decline and disability, which are commonly precipitated by intervening illnesses and injuries.^26,27^ Second, disadvantaged neighborhoods may be less walkable because of the absence or poor maintenance of sidewalks or concerns about personal safety, reducing participation in structured physical activity that has been shown to prevent disability.^28^ Third, other healthy habits may be more difficult to maintain in disadvantaged neighborhoods because of limited access to affordable and nutritious food, leading to or exacerbating diet-related chronic diseases and obesity.^29^ Fourth, many acute and chronic conditions may be triggered or worsened by mental stress associated with living in neighborhoods with heightened levels of crime and violence.^30^ Fifth, disadvantaged neighborhoods often lack public transportation infrastructure and access to formal social resources that may leave older persons with unmet needs, making them more susceptible to the onset and progression of disability.^31^

Addressing these and other disparities in resource-poor environments will require substantial investments to support a more robust set of social and public health interventions.^30,32^ Based on the accumulating evidence linking socioeconomically disadvantaged neighborhoods to an increasing number of adverse health outcomes,^1-4^ these investments could potentially lead to cost savings through large-scale reduction of inequalities in morbidity and mortality.^32,33^ The outcomes evaluated in the current study, active and disabled life expectancy, are important not only to policy-makers, but also to older persons who consistently identify the maintenance of independent function as their top health outcome priority.^34^ Our findings that living in disadvantaged neighborhoods is associated with lower active life expectancy and a greater percentage of projected remaining life with disability suggest that new or expanded social and public health initiatives could also improve the functional well-being of older persons.

As in prior studies,^2,35^ ADI scores were dichotomized to distinguish the disadvantaged from non-disadvantaged groups. This practice conforms with fundamental theories of social disadvantage,^36^ which suggest that persons can compensate up to a certain threshold beyond which additional disadvantage leads to adverse outcomes.^37^ Because our objective was to evaluate neighborhood disadvantage as a contextual factor rather than as a proxy for individual disadvantage, our findings are not susceptible to ecological fallacy, in which a region’s aggregate traits are inappropriately attributed to a particular person.^38^ An important strength of the current study is the ability to account for individual-level socioeconomic characteristics and other prognostic factors. The reductions observed in disabled life expectancy among participants from disadvantaged neighborhoods in the adjusted analyses, which diminished the non-significant differences between the disadvantaged and non-disadvantaged groups in the unadjusted analyses, were likely attributable to the higher prevalence of several individual-level factors that conferred increased risk for disability in the disadvantaged group, including female sex, lower education and income, and cognitive impairment.

Additional strengths of the study include monthly assessments of disability with little missing data and long duration of follow-up with relatively little attrition for reasons other than death. The availability of monthly data allowed us to account for high rates of transitions between states of disability and independence^39^ and should lead to estimates of active and disabled life expectancy that are more accurate than those based on less frequent assessments.

Our study has several limitations. First, because this was an observational study, the reported associations cannot be construed as causal relationships. Second, although our multivariable analyses accounted for many individual-level socioeconomic characteristics and other prognostic factors, our findings could be confounded by unmeasured factors that are associated with both neighborhood disadvantage and active/disabled life expectancy. Third, because our study participants were members of a single health plan in South Central Connecticut, our results may not be generalizable to older persons in other settings. The generalizability of our results is enhanced by our high participation rate, which was greater than 75%, and low rate of attrition.^40^ Finally, participants in disadvantaged neighborhoods were identified based on the worst quintile of ADI scores for Connecticut, corresponding approximately to the worst two quarters of ADI scores for the entire US. The differences reported in the current study may be more marked in national samples of community-living older persons.

In summary, living in a disadvantaged neighborhood is associated with lower active and total life expectancy and a greater percentage of projected remaining life with disability. By addressing deficiencies in resource-poor environments, new or expanded social and public health initiatives have the potential to improve the functional well-being and longevity of community-living older persons and, in turn, reduce health disparities in the US.

## Data Availability

Dr. Gill had full access to all of the data in the study and takes responsibility for the integrity of the data and the accuracy of the data analysis.

## Acknowledgments

We thank Denise Shepard, BSN, MBA, Andrea Benjamin, BSN, Barbara Foster, and Amy Shelton, MPH, for assistance with data collection; Geraldine Hawthorne, BS, for assistance with data management; Peter Charpentier, MPH, for design and development of the study database and participant tracking system; and Joanne McGloin, MDiv, MBA, for leadership and advice as the Project Director. Each of these persons were paid employees of Yale School of Medicine during the conduct of this study.

## Author Contributions

Dr. Gill had full access to all of the data in the study and takes responsibility for the integrity of the data and the accuracy of the data analysis. The specific contributions are enumerated in the authorship, financial disclosure, and copyright transfer form.

## SUPPLEMENTARY APPENDIX

**Table S1**. Indicators of the Area Deprivation Index at the Census-Block Level

