## Supplementary material for "Neighborhood Disadvantage and Active, Disabled and Total Life Expectancy Among Community-living Older Persons": Table S1

**Table S1.** Indicators of the Area Deprivation Index at the Census-Block Level

##### 1. Percentage of housing units without complete plumbing

##### 2. Percentage population aged ≥25 y with <9 years of education

##### 3. Percentage population aged ≥25 y with <12 years of education

##### 4. Percentage of employed persons aged ≥16 years with white collar occupation

##### 5. Median family income

##### 6. Income disparity

##### 7. Percentage civilian labor force aged ≥16 years

##### 8. Percentage of households with >1 person per room

##### 9. Median home value

##### 10. Median gross rent

##### 11. Median monthly mortgage

##### 12. Percentage occupied housing units

##### 13. Percentage families below the poverty level

##### 14. Percentage population <150% of poverty threshold

##### 15. Percentage of single‐parent household with children aged <18 years

##### 16. Percentage of households without motor vehicle

##### 17. Percentage of households without telephone
